# Performance characteristics of a rapid SARS-CoV-2 antigen detection assay at a public plaza testing site in San Francisco

**DOI:** 10.1101/2020.11.02.20223891

**Authors:** Genay Pilarowski, Paul Lebel, Sara Sunshine, Jamin Liu, Emily Crawford, Carina Marquez, Luis Rubio, Gabriel Chamie, Jackie Martinez, James Peng, Douglas Black, Wesley Wu, John Pak, Matthew T. Laurie, Diane Jones, Steve Miller, Jon Jacobo, Susana Rojas, Susy Rojas, Robert Nakamura, Valerie Tulier-Laiwa, Maya Petersen, Diane V. Havlir, The CLIAHUB Consortium, Joseph DeRisi

## Abstract

We evaluated the performance of the Abbott BinaxNOW™ Covid-19 rapid antigen test to detect virus among persons, regardless of symptoms, at a public plaza site of ongoing community transmission. Titration with cultured clinical SARS-CoV-2 yielded a human observable threshold between 1.6×10^4^-4.3×10^4^ viral RNA copies (cycle threshold (Ct) of 30.3-28.8 in this assay). Among 878 subjects tested, 3% (26/878) were positive by RT-PCR, of which 15/26 had a Ct<30, indicating high viral load. 40% (6/15) of Ct<30 were asymptomatic. Using this Ct<30 threshold for Binax-CoV2 evaluation, the sensitivity of the Binax-CoV2 was 93.3% (14/15), 95% CI: 68.1-99.8%, and the specificity was 99.9% (855/856), 95% CI: 99.4-99.9%.

## INTRODUCTION

The global pandemic of SARS-CoV-2 infection has spread at an unprecedented pace [1]. Efficient transmission of infection by the respiratory route, including by asymptomatic and pre-symptomatic persons, has fueled ongoing surges of new cases. Countries that have succeeded in epidemic control have done so through the use of non-pharmacologic measures to reduce transmission (universal masking, social distancing) coupled with aggressive testing, tracing and isolation of infected subjects and quarantine of their contacts [2,3].

To date, the cornerstone of testing has been quantitative RT-PCR examination of respiratory secretions. Such testing has excellent sensitivity and specificity but is expensive and requires sophisticated equipment and highly trained personnel to complete [4]. In practice in the United States, these features have often generated testing bottlenecks that can result in long delays in the reporting of results, compromising their utility in epidemiologic investigation and high frequency of testing approaches [5]. As a result, there is increasing interest in more rapid and economical assays that circumvent these limitations [6]. However, methods that do not include an amplification step are inherently less sensitive; thus, their proper deployment in populations will therefore require a rigorous evaluation of their performance characteristics in the different epidemiologic settings in which they may be used.

Lateral flow antigen detection diagnostics have long been deployed for a variety of infectious diseases including malaria, RSV, influenza, and more. For the current pandemic, one of the most available of these tests is the Abbott BinaxNOW™ COVID-19 Ag Card (hereafter referred to as Binax-CoV2), which detects viral nucleocapsid (N) protein in direct nasal swabs. The test requires no laboratory instrumentation; results are scored visually and returned within 15 minutes. In August 2020, the FDA issued an emergency use authorization (EUA) for this test in the diagnosis of SARS-CoV-2 infection in symptomatic patients being tested within 7 days of symptom onset [7]. The US Department of Health and Human Services is now distributing 150 million test kits to the 50 states, including some directly to nursing homes, assisted living facilities and other high-risk settings; allocation of the remaining tests will be decided by state governments. Given the value of a rapid assessment of infectiousness, there is anticipated use in a broad range of subjects including those who are asymptomatic.

Here we present the first systematic examination of the performance characteristics of the Binax-CoV2 test in a community screening setting where testing was offered for symptomatic and asymptomatic subjects. Additionally, we present data from controlled laboratory evaluations of the Binax-CoV2 test and quantification of inter-lot variability.

## METHODS

### Study population and Specimen Collection

Over 3 days in September 2020, we offered testing in the Mission District, a Latinx-predominant neighborhood, known from prior surveys to have an elevated prevalence of SARS-CoV-2 infection [8,9]. Walk-up, free outdoor testing was conducted at a public plaza located at a point of intersection between the Bay Area-wide subway system (BART) and the San Francisco city bus/streetcar system (MUNI). On the day of test, participants self-reported symptoms and date of onset, demographics, and contact information, as required by state and federal reporting guidelines. On each participant, a laboratory technician performed sequential anterior swab (both nares per swab) for the Binax-CoV2 assay followed by a second swab (both nares) for RT-PCR. Participants were notified of RT-PCR test results via standard procedures; because of uncertainties of testing performance in this population and setting, Binax-CoV2 results were not reported back to study subjects.

### Laboratory Testing for SARS-CoV-2

RT-PCR detection of SARS-CoV-2 was performed at the CLIA-certified lab operated by UCSF and the Chan Zuckerberg Biohub as described [10,11]. Briefly, anterior nasal swabs collected in DNA/RNA Shield (Zymo Research) were subjected to RT-PCR using probes specific to the viral N and E genes, and to an internal human positive control (RNAse P). The assay has a detection limit of 100 viral copies/mL, and a sample is designated as positive if either the N or E probes yield cycle thresholds of less than 45.

### Field Testing using Binax-CoV2 assay

Binax-CoV2 assay was performed by technicians on site as described by the manufacturer using the supplied Puritan swabs. Each assay was read by two independent observers, and a third observer served as a “tie-breaker”. Beginning on day 2 of the study, each Binax-CoV2 assay card was scanned onsite using a color document scanner (CanoScan LIDE 400, Canon). For the purpose of this paper, the sample bands were retrospectively quantified from image data. Briefly, sample and background regions were localized by offset from the control band, and relative mean pixel intensity decreases were calculated from blue and green channels averaged with respect to background.

### Titration of in vitro cultured SARS-CoV-2 on Binax-CoV2 cards

SARS-CoV-2 from a UCSF clinical specimen was isolated, propagated and plaqued on Huh7.5.1 cells overexpressing ACE2 and TMPRSS2 [12]. Viral titers were determined using standard plaque assays with Avicel RC-591 [13]. For viral titration experiments, SARS-CoV-2 with known titer was diluted in Dulbecco’s phosphate-buffered saline (DPBS) and 40 microliters of each dilution was absorbed onto Puritan Sterile Foam Tipped Applicator swabs. After absorption, antigen detection was completed using Binax-CoV2 per manufacturer instructions. Images of Binax-CoV2 cards were taken on an Apple iPhone6. Each dilution was also assayed by RT-PCR calibrated with internal cloned viral RNA standards. All experiments using cultured SARS-CoV-2 were conducted in a biosafety level 3 laboratory.

### N protein titration assay of Binax-CoV2 lots

SARS-CoV-2 N protein (1-419) was expressed in BL21(DE3) *E*.*coli* and purified by Ni-NTA chromatography, incorporating a 1M NaCl, 50 mM imidazole wash to remove bound RNA, and formulated in 50 mM Na2HPO4, 500 mM NaCl, 10% glycerol, pH 8.0 at 1.72 mg/mL. Purified protein was diluted in 50 mM Na2HPO4, 500 mM NaCl, pH 7.2. Six concentrations of N protein were tested on ten lots of Binax-CoV2 kits: 9 lots obtained from the State of California and 1 original lot used in the community study and laboratory live virus work (126029). Briefly, 40ul of purified N protein was absorbed onto the provided Puritan swab. Binax-CoV2 card tests were run per manufacturer instructions by two technicians per lot for a total of four replicates per concentration and imaged on document scanner. The Abbott-provided positive control swab from each lot was run and passed quality control for all ten lots.

### Ethics statement

The UCSF Committee on Human Research determined that the study met criteria for public health surveillance. All participants provided informed consent for dual testing.

## RESULTS

### Binax-CoV2 performance using a titration of in vitro cultured SARS-CoV-2

To explore the relationship of RT-PCR cycle threshold (Ct), viral load, and the corresponding visual Binax-CoV2 result, a dilution series of lab-cultured SARS-CoV-2 with known titers was assayed both by RT-PCR and by Binax-CoV2 (**Figure 1**). For this stock of virus, the threshold for detectability by human eye on the Binax-CoV2 assay was between 1.6-4.3×10^4^ viral copies (100-250 pfu), corresponding to a Ct (average of N and E genes) of 30.3 and 28.8, respectively in this assay.

**Figure 1.**
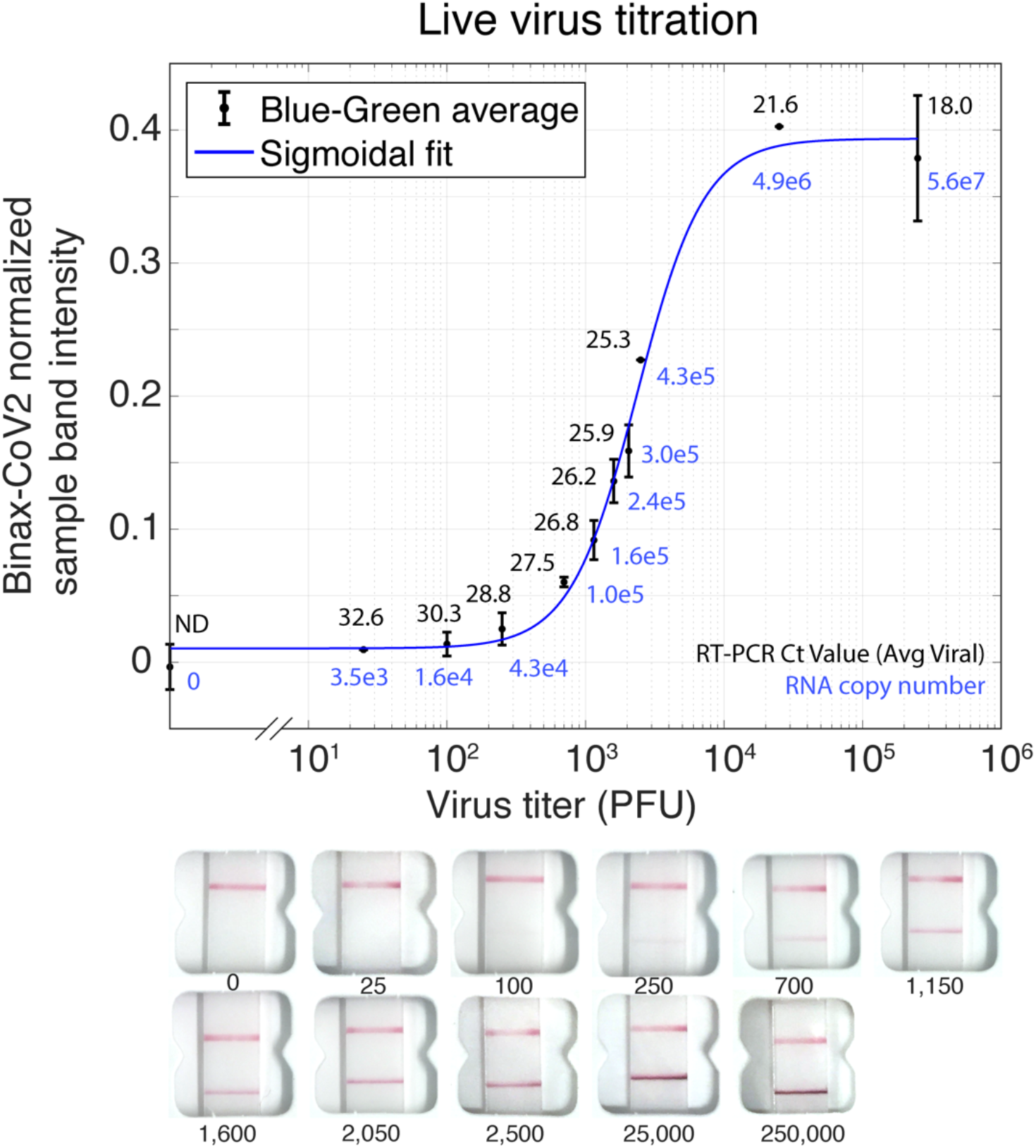
Titration of in vitro grown SARS-CoV-2 and detection on Binax-CoV2 assay. Normalized Binax-CoV2 sample band intensity (blue-green average) for cards loaded with a known amount of virus. Error bars represent standard deviation of sample band intensity of technical replicates. RT-PCR testing was performed at the CLIAHUB consortium [10]. Corresponding RT-PCR Ct values (average of N and E gene probes) are printed in black and the corresponding RNA copy number printed in blue. Note that Ct and genome copy number correlation varies by RT-PCR platform. Representative card images from each datapoint are shown below.

### Community RT-PCR Testing Results

Of the 878 subjects tested, 54% were male, 77% were 18 to 50 years of age, 81% self-identified as Latinx, and 84% reported no symptoms in the 14 days before testing. Twenty-six persons (3%) were RT-PCR+; of these, 15/26 (58%) had a Ct<30 and 6/15 (40%) were asymptomatic. Among asymptomatic individuals with Ct<30, 4/6 developed symptoms within 2 days after testing. Of the 11 persons RT-PCR-positive with Ct>30, 4 reported symptom onset ≥ 7 days before testing, 1 reported symptom onset 3 days prior to testing, and the remainder reported no symptoms.

### Comparison of RT-PCR and Binax-CoV2 testing results from Community Testing

Because the readout of the Binax-CoV2 assay is by visual inspection of the bands on the lateral flow assay strip, there is an element of subjectivity in scoring the results, especially when bands are faint or partial (i.e. do not extend across the entire width of the strip). The manufacturer’s instructions suggest scoring any visible band (partial or full, faint or strong) as positive. Because these criteria were elaborated from studies of symptomatic cases, we were uncertain of their applicability to the screening of populations with asymptomatic subjects. Accordingly, we used the manufacturer’s reading instructions and tested 217 samples, of which 214 yielded valid Binax-CoV2 results: 7 (3.3%) were RT-PCR (+); using the manufacturer’s proposed criteria, 5 of these 7 were Binax-CoV2 (+). However, of 207 RT-PCR (-) samples, 9 (4.3%) were Binax-CoV2 (+). Thus, using the manufacturer’s proposed criteria, 9/14 Binax-CoV2 (+) tests (64%) in this population were likely false positives (Ag(+)/RT-PCR(-)). Clearly, these initial criteria were problematic in a screening setting like this one.

Therefore, on subsequent test days, we evaluated additional criteria for classifying a band as positive, in consultation with experts from the manufacturer’s research staff. Classifying only strong bands as positive eliminated false positives, but did not address the subjective thresholding process, particularly for calling faint bands. Optimal performance occurred the when bands were scored as positive *if they extended across the full width of the strip, irrespective of the intensity of the band*. Using these criteria on 283 RT-PCR-negative samples, none scored positive for antigen on the Binax-CoV2 test, thus markedly alleviating the false positive readings. With these updated scoring criteria, 5/9 RT-PCR (+) samples were Binax-CoV2 (+) for antigen. The 4/9 RT-PCR (+) samples that were Binax-CoV2 (-) had Ct>30. We find that scoring a test as positive if bands extended across the full width of the strip, irrespective of band intensity, the least subjective and easiest method to implement in the field and have developed a training tool: https://unitedinhealth.org/binax-training. Accordingly, this method was used to score the data collected in this study (**Figure 2**).

**Figure 2.**
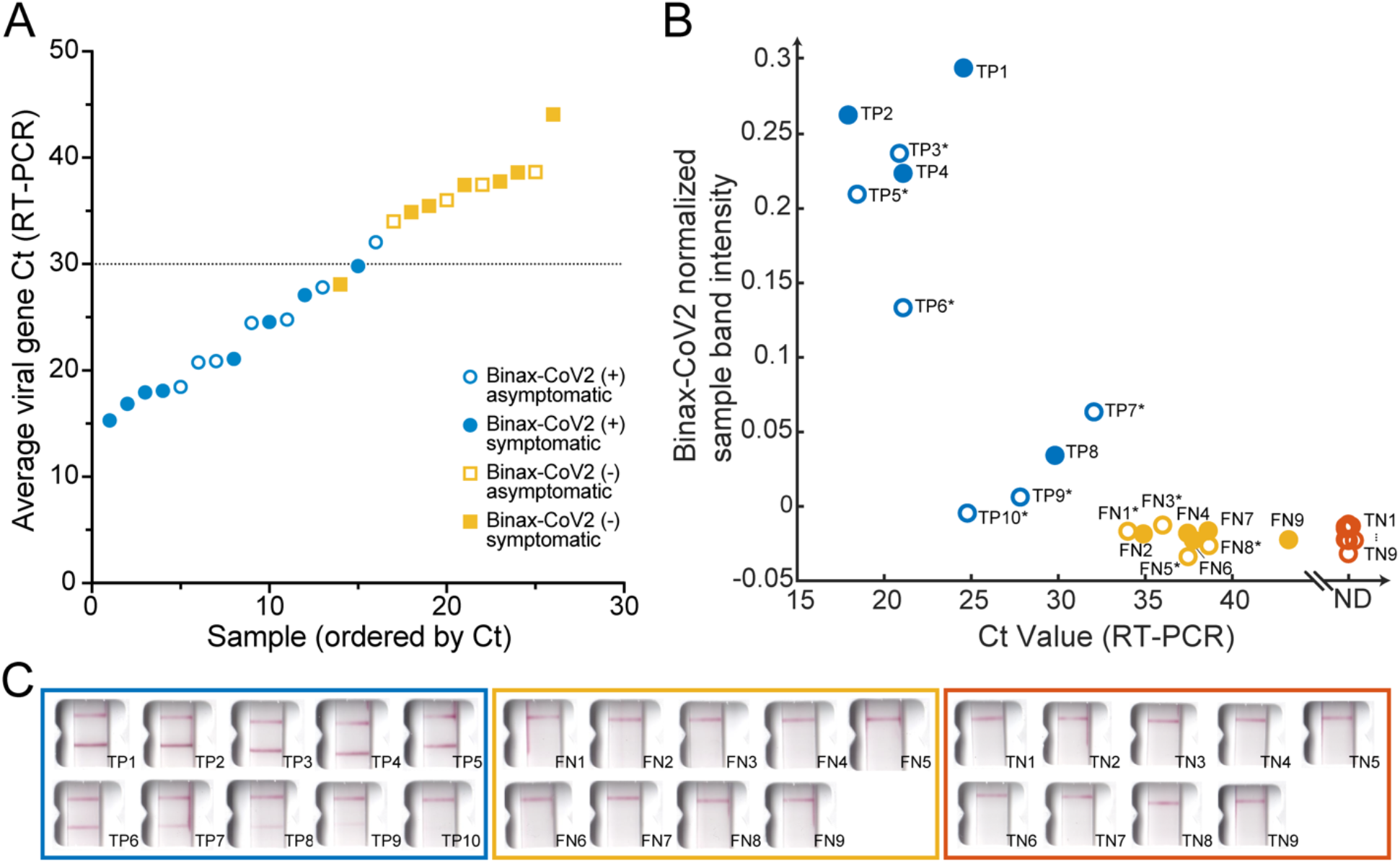
Comparison of Binax-CoV2 test with quantitative RT-PCR test. (**A**) Average viral Ct values from all 26 RT-PCR-positive individuals from the community study are plotted in ascending order. Blue circles indicate Binax-CoV2-positive samples and yellow squares indicate Binax-CoV2-negative samples. Empty symbols represent individuals who were asymptomatic on day of test and filled symbols represent individuals who reported symptoms on day of test. (**B**) Normalized sample band signal from retrospective image analysis of Binax-CoV2 cards was plotted as a function of Ct value for all available scanner images (19/26 RT-PCR positives and a random subset of RT-PCR negatives). Binax-CoV2 True Positives are shown in blue with ‘TP’ labels, False Negatives in yellow with ‘FN’ labels, and True Negatives in red with ‘TN’ labels. (**C**) Corresponding Binax-CoV2 card images from the data in panel B.

The Binax-CoV2 assay results of the 26 RT-PCR-positive individuals are stratified by the Ct value of the RT-PCR test and shown in **Figure 2**. As might be expected, the rapid antigen detection test performed well in samples with higher viral loads: 15 of 16 samples with Ct<32 were positive (**Figure 2a**). By contrast, none of the 10 samples with Ct≥34 were positive by Binax-CoV2 antigen detection. Retrospective image quantification of Binax-CoV2 sample bands correlates with RT-PCR Ct values for those individuals (**Figure 2b**). In each case, the corresponding image is shown in order to demonstrate the correspondence between RT-PCR and the visual result (**Figure 2c**).

### Sensitivity and Specificity

RT-PCR is considered the gold standard for SARS-CoV-2 detection [4] and, in this assay, has a limit of detection of 100 viral RNA copies per mL. Direct antigen assays, such as Binax-CoV2, are unlikely to rival the sensitivity of RT-PCR. Thus, to quantify the performance on the Binax-CoV2 assay in the context of community based testing, we defined a threshold for high virus levels corresponding to the range thought to be the most transmissible: a cycle threshold of 30, which corresponds to a viral RNA copy number of approximately 1.9×10^4^ in this assay [11]. Using this Ct<30 case definition and 95% confidence interval (CI), the sensitivity of the Binax-CoV2 was 93.3%, CI: 68.1-99.8% (14/15), and the specificity was 99.9%, CI: 99.4-99.9% (855/856). Adjusting the threshold to a more conservative cycle threshold value of 33 (2.6×10^3^ viral RNA copies), the sensitivity was 93.8%, CI: 69.8-99.8% (15/16) and the specificity was 100%, CI: 99.6-100% (855/855).

### Evaluation of Binax-CoV2 lot-to-lot variation

We quantified lot to lot variability in 10 different lots of Binax-CoV2 card tests using a dilution series of N protein, the SARS-CoV-2 protein that is captured in the Binax-CoV2 assay (**Supplementary Figure 1**). At protein concentrations 17.2ng/ml and greater, sample band was detected in all lots, and thus would not affect the outcome of this binary assay (**Supplementary Figure 1A**). The binary scoring of the Binax-CoV2 tests is likely to be affected by lot to lot variability only at lower levels of viral protein concentration, near the inherent limit of detection of the test.

## DISCUSSION

These data provide the first quantitative analysis of the performance characteristics of a widely disseminated rapid antigen detection kit when applied to a population-based cohort that included asymptomatic subjects. These results indicate a clear relationship between relative viral load and test positivity, and provide a practical, real-world criterion to assist calling results in this setting. We found that small modifications to the training of Binax-CoV2 technicians reduced the presence of false-positives, a legitimate concern for the roll-out of these tests. Importantly, directing technicians to call positives only if the sample band extended edge-to-edge of the test strip appeared to preserve sensitivity while also reducing false-positives results.

The currently approved EUA for the Binax-CoV2 assay specifies use only in symptomatic individuals. One of the main drivers of the current pandemic is that a substantial percentage of infected people do not report symptoms, despite having viral loads indistinguishable from symptomatic individuals [8,14]. The results presented here suggest that the Binax-CoV2 test should not be limited to symptomatic testing alone but should also incorporate asymptomatic individuals. Limiting use of Binax-CoV2 to symptomatic individuals would have missed nearly half of the SARS-CoV-2 infections with high viral loads.

Furthermore, the impact of effectively deployed tests on forward transmission is hampered by long wait times, often days, for test results. We reported previously that in the community setting, by the time a person is tested, counseled and situated under effective isolation conditions, the effective isolation period is often nearly over [9]. This is particularly true for many communities of color, where reported delays in accessing tests and results are even longer [5,15]. Rapid tests could reduce these delays and maximize time of effective isolation. Limitations of our study include its cross-sectional design and overall small number of RT-PCR positive cases. Additional field performance of this assay is needed and will help inform optimal use strategies. We recommend evaluating the Binax-CoV2 assay side by side with RT-PCR in each context it will be used prior to use of Binax-CoV2 without the use of RT-PCR.

During the early stages of infection, viral load may be too low to detect by direct antigen assays, such as Binax-CoV2. This inherent lower sensitivity may be offset by faster turn-around, the ability to test more frequently, and overall lower cost, relative to traditional RT-PCR methods. That said, for persons who present with a high index of suspicion of COVID-19 and a negative Binax-CoV2 result, the test should be complimented with RT-PCR or a repeat Binax-CoV2 test at a later time to make sure case not missed.

In summary, under field conditions with supplementary technician training, the Binax-CoV2 assay accurately detected SARS-CoV-2 infection with high viral loads in both asymptomatic and symptomatic individuals. The Binax-CoV2 test could be a valuable asset in an arsenal of testing tools for the mitigation of SARS-CoV-2 spread, as rapid identification of highly infectious individuals is critical.

## Data Availability

All data referred to in the manuscript is included in the figures of the manuscript. Supplemental material (Binax-CoV2 reader training guide) is included in the link.

https://unitedinhealth.org/binax-training

## ACKNOWLEDGMENTS

We thank Bevan Dufty and the BART team, Jeff Tumlin and the San Francisco MUNI, Supervisor Hillary Ronen, Mayor London Breed, Dr. Grant Colfax and the Department of Public Health, Salu Ribeiro and Bay Area Phlebotomy and Laboratory services, PrimaryBio COVID testing platform, and our community ambassadors and volunteers. We would like to thank Dr. Don Ganem for his writing assistance and critical discussion, Dr. Andreas Puschnik for Huh7.5.1 overexpression cells used for SARS-CoV-2 growth, and Drs. Terry Robins, Stephen Kovacs, and John Hackett Jr from Abbott Laboratories for their support. We would also like to thank both Abbott Laboratories and the California Department of Public Health for their generous donations of BinaxNOW™ COVID-19 Ag cards.

## FIGURES

**Supplementary Figure 1.**
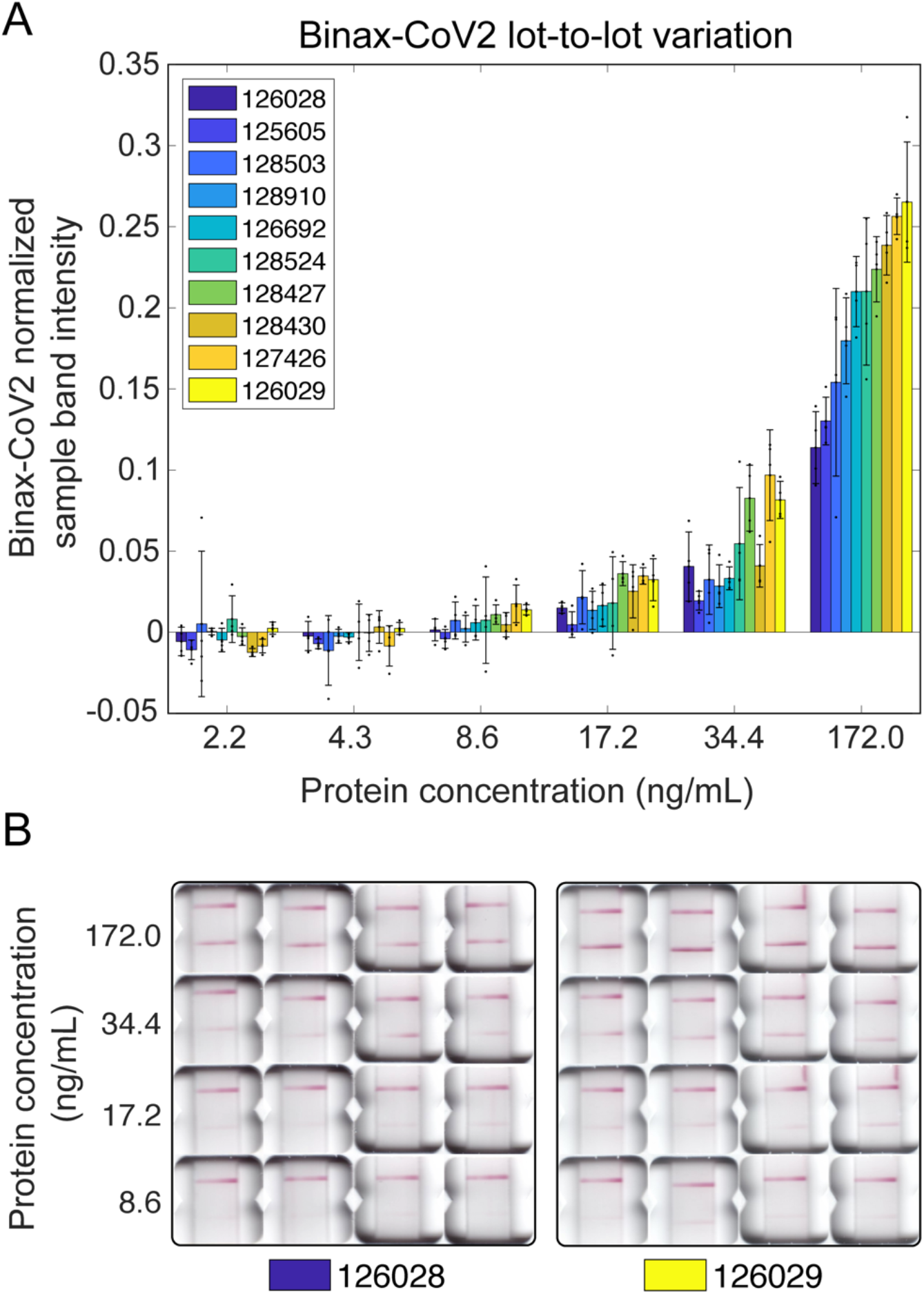
Variability of signal intensity in Binax-CoV2 card lots. (**A**) Normalized sample band signal intensity of Binax-CoV2 cards from different lots run with a dilution series of purified SARS-CoV-2 N protein with known concentration. N=4 cards per lot per concentration. Each point represents one card. (**B**) Images of each card test for the highest (126029) and lowest (126028) performing lots.

